# Management of malaria in children under 5-years-old during COVID-19 pandemic in Sierra Leone: a lesson learned?

**DOI:** 10.1101/2020.11.04.20225714

**Authors:** Danilo Buonsenso, Francesco Iodice, Bianca Cinicola, Francesca Raffaelli, Solia Sowa, Walter Ricciardi

## Abstract

Growing evidences are showing the potential indirect effects of COVID-19 on the health systems of low-resource settings, where diseases such as Tuberculosis, HIV and Malaria represent major killers. Therefore, we performed a retrospective study aimed to evaluate the impact of COVID-19 on Malaria programs in a peripheral region of Sierra Leone, previously involved by the Ebola outbreak in 2015, when malaria care have been impaired since local health systems were overwhelmed by Ebola cases. During COVID-19 in Sierra Leone, we did not notice a drop in malaria diagnosis in children, suggesting that a proactive approach in the management of malaria in endemic countries during COVID-19 may have had a positive impact. A comprehensive approach that include also educational activities to sensitize the local population, was useful to guarantee successful malaria diagnosis and treatment, and prevents excess of malaria deaths due to potential disruption of the local health systems related to the SARS-CoV-2 pandemic.

**Contribution to the field:** we performed a retrospective study aimed to evaluate the impact of COVID-19 on Malaria programs in a peripheral region of Sierra Leone, previously involved by the Ebola outbreak in 2015, when malaria care have been impaired since local health systems were overwhelmed by Ebola cases. During COVID-19 in Sierra Leone, we did not notice a significant change in the middle term period in malaria diagnosis in children, suggesting that a proactive approach in the management of malaria in endemic countries during COVID-19 may have had a positive impact. A comprehensive approach that include also educational activities to sensitize the local population, was useful to guarantee successful malaria diagnosis and treatment, and prevents excess of malaria deaths due to potential disruption of the local health systems related to the SARS-CoV-2 pandemic.

**Funding statement:** nothing to declare

**Ethics statements:** *Studies involving animal subjects:* Generated Statement: No animal studies are presented in this manuscript.

*Studies involving human subjects:* Generated Statement: The studies involving human participants were reviewed and approved by Bureh Town Community Hospital. Written informed consent to participate in this study was provided by the participants’ legal guardian/next of kin.

*Inclusion of identifiable human data:* Generated Statement: No potentially identifiable human images or data is presented in this study.

*Data availability statement:* Generated Statement: The raw data supporting the conclusions of this article will be made available by the authors, without undue reservation.

## Introduction

After the first description in China, SARS-CoV-2 spread all over the world and reached sub-Saharan Africa. Even in this part of the world, the number of COVID-19 cases and deaths are rising day by day, although not on a significant rate compared with Europe, Asia, North and South America. The reasons of such a difference are still unexplained, but multiple factors can be involved, including genetic differences and lack of testing points in most areas, particularly in the peripheries.

Anyway, the arrival and continuous spread of SARS-CoV-2 in Africa is worrying major organiaztions (including the World Health Organization and major medical journals) because of the direct impact of the viral spread on a weak health system and the indirect effects of lockdown in a weak economic system (1). Preliminary data arrived from Sierra Leone where we previously highlighted the social consequences of SARS-CoV-2 lockdown (2). We reported how restrictive measures had a strong impact on the population with majority of people who live in peripheries are losing their jobs and householders, the people to whom most of the family’s financial support is assigned, having serious concerns in providing basic needs (water and food) to their relatives.

However, it was immediately clear that a special attention of the health system should be paid to the possible effects of COVID-19 pandemic on other major killers in Africa: HIV, tuberculosis and Malaria. During the last decade, African countries greatly improved the care of patients with these conditions, thanks to a coordinated approach including governments acts, the support of major international agencies (such as the World Health Organization (WHO) and non-governmental organizations (NGO). However, the COVID-19 pandemic could possibly break a decade of successes in tuberculosis, HIV and Malaria care. The impact on tuberculosis and HIV care has been widely claimed (3) and we showed how the number of suspected and confirmed tuberculosis cases dropped in an outpatient unit in Sierra Leone (4). The estimated impact of these measures on tuberculosis care could be considered impressive, with major organizations estimating millions of new cases in the next years due to missed diagnosis or incomplete treatments (and subsequent community spread) during COVID-19 lockdown (https://www.theunion.org/news-centre/news/union-warns-the-covid-19-pandemic-must-not-divert-attention-from-the-needs-of-children-and-adolescents-in-tb-endemic-african-countries).

Importantly, there is the problem of stigmatization: people are worried to be recognized as COVID-19 patients and health center has been improperly recognized as potential spots where the contagion would be easier (4). During Ebola Virus Disease (EVD) outbreak, several violent riots happened at health centers across the most involved countries, moved by the believe that medical staff were somehow responsible for spreading EVD to the villages. This resulted in further disruption of the control programs (5).

During the World Malaria Day on April 25^th^, fears about potential interruption of Malaria control programs in endemic countries due to COVID-19 have been raised (6). Most of these fears come from the consequences of EVD on malaria care in 2014-2016, when the number of malaria diagnoses, the number of patients with malaria seeking appropriate health care and the volume of malaria treatments being dispensed dropped significantly compared with previous years (6-8). It was estimated that about 7000 additional malaria-associated deaths among children younger than 5 years in Guinea, Liberia, and Sierra Leone due to the Ebola outbreak (6, 9).

In the case of Ebola, clinical similarity of EVD with malaria, fear of contracting Ebola in the health-care facilities, and interruption of distribution of insecticide-treated bed-nets (ITNs) were all contributing factors leading to disruption of malaria care (10).

There is now the real possibility that COVID-19 could have the same indirect impact on Malaria. Considering that Malaria mainly kills children under five years of age, while COVID-19 is relatively mild in this group (11), the potential effects of disrupted malaria programs on children’s health could be massive. Therefore, we performed this retrospective study aimed to evaluate the impact of COVID-19 on Malaria programs in a peripheral region of Sierra Leone, previously involved by the EVD outbreak in 2015.

## Methods

To understand the impact of COVID-19 on Malaria care, we retrospectively evaluated the gross numbers of children aged 0 to 60 months diagnosed with confirmed *P. falciparum* Malaria in the Konta Wallah Community Health Center of Port Loko Discrtict, Kamasondo Chiefdom, a government recognized Malaria inpatient and outpatient unit, referral for an area of about 8,000 people. In this setting, malaria is diagnosed either on a clinical bases or confirmed with an antigen rapid test able to detect *P. falciparum* antigen. We collected the number of patients tested with confirmed *P. falciparum* Malaria during the first 5 months of the year 2020 (January, February, March, April, May), and compared it with the cases reported in 2018 and 2019 during the same months. The study was approved by a local commission composed of the responsible of the Malaria unit of the local health centre (S.S.), the research team of the Bureh Town Community Hospital of Sierra Leone, the headman of the community and the old men of the village, in a similar way to what happens for all the important political and economic decisions in the examined area (n25_may-2020). Personal data were not collected. In Sierra Leone, the first COVID-19 presumptive cases have been documented at the end of March 2020 and lockdown declared in April 2020. On May 2020, the lockdown has been retired but people were not allowed to move between districts. Descriptive and statistical analyses performed with STATA v16. Dataset available upon reasonable request contacting the corresponding author.

## Results

During the period analysed, fluctuations in Malaria diagnoses have been documented (figure 1) and, although on April 2020 there has been reduction in diagnosed cases compared with March 2020 and April 2019 (P 0.01), the number of cases did not differ from April 2018 (P > 0.05, figure 1a and 1b). On May 2020 a new raise in malaria diagnoses was found, compared with April 2020, but comparable to May 2019 and 2018 (P > 0.05, figure 1c). The number of malaria deaths in children younger than 5 years of age was similar in 2018 and 2019, and 2 cases detected from January to May 2020 (potentially a similar rate/year considering the short timeframe evaluated in 2020) (figure 1d).

**Figure 1.**
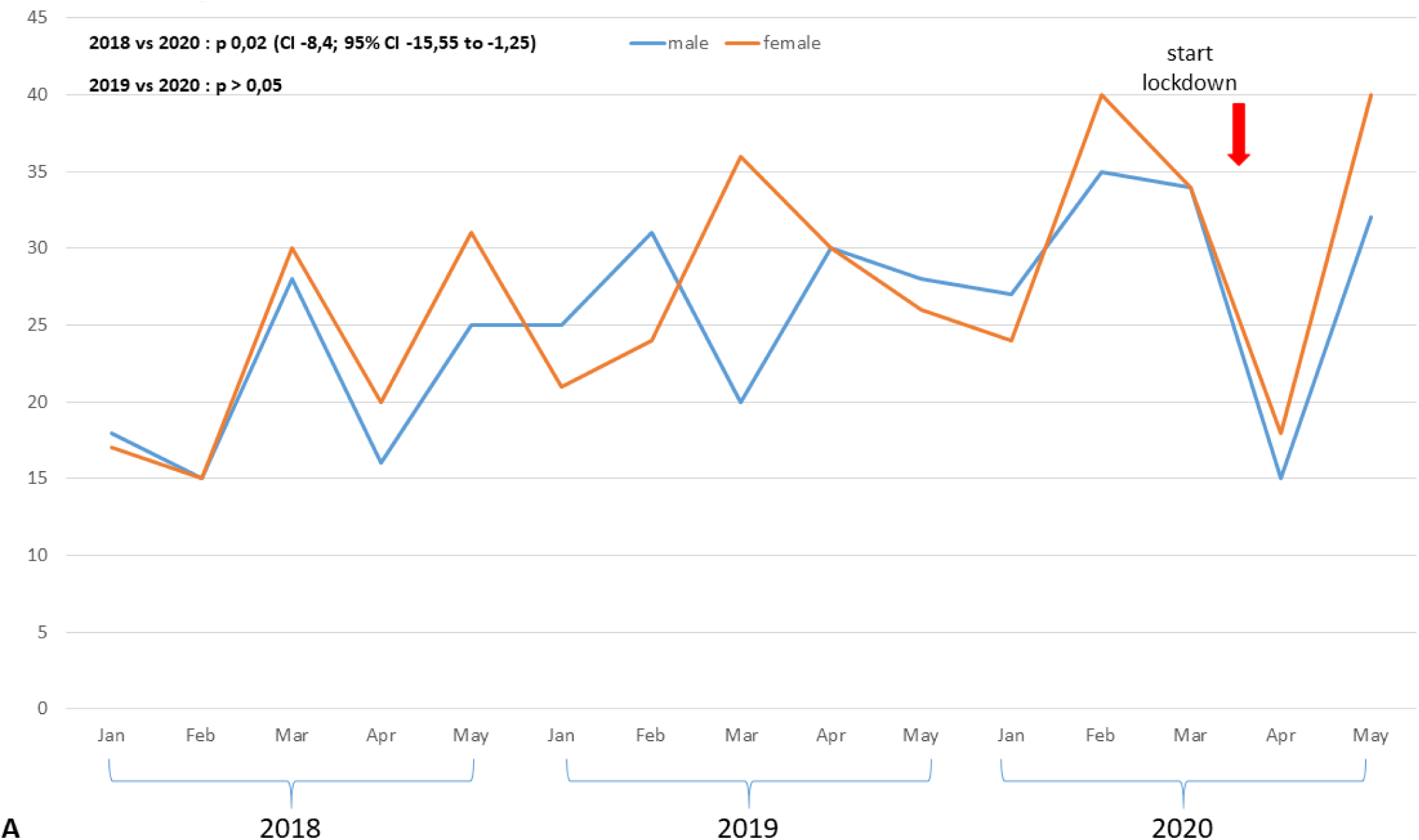

**Figure 2.**
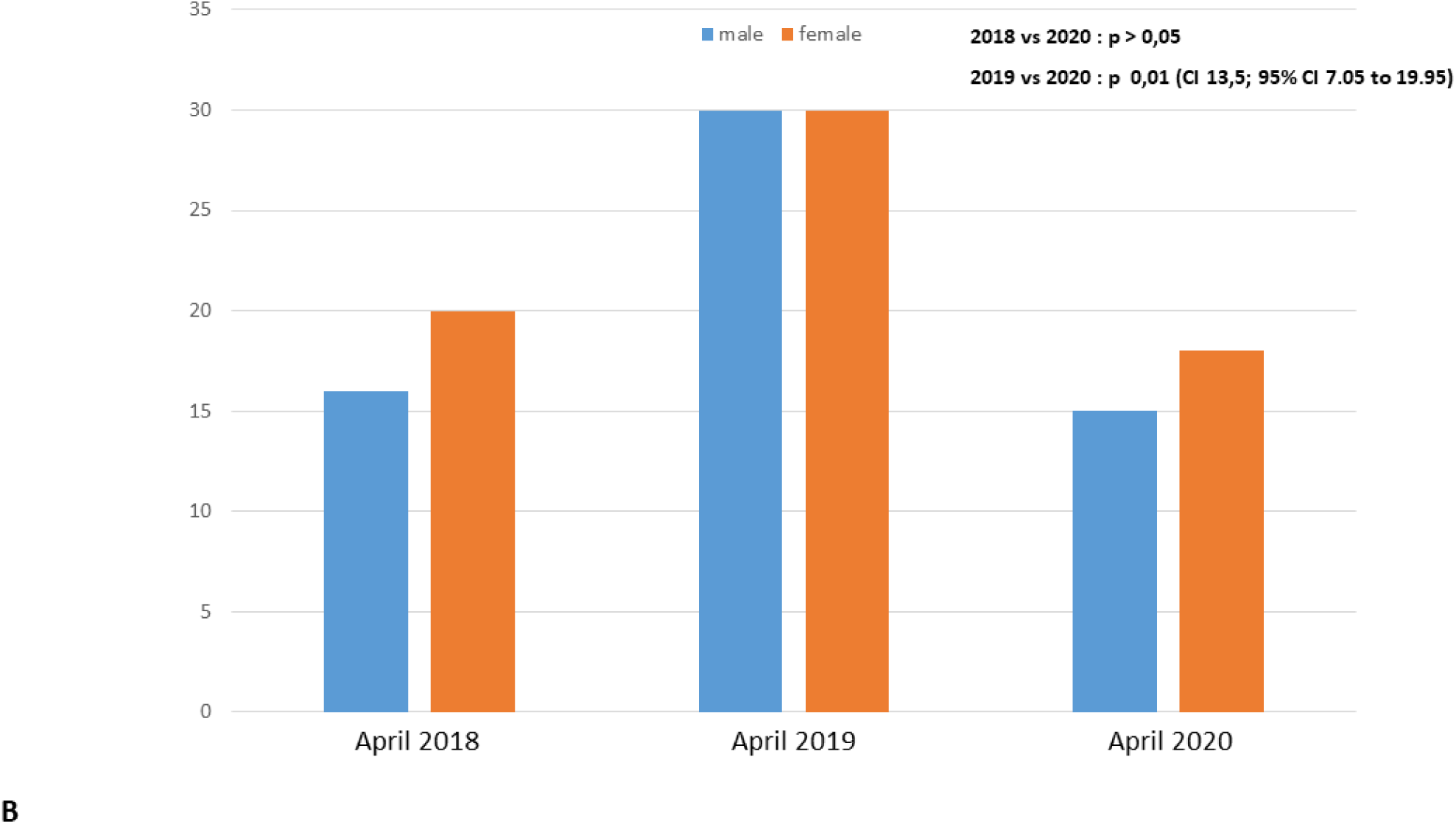

**Figure 3.**
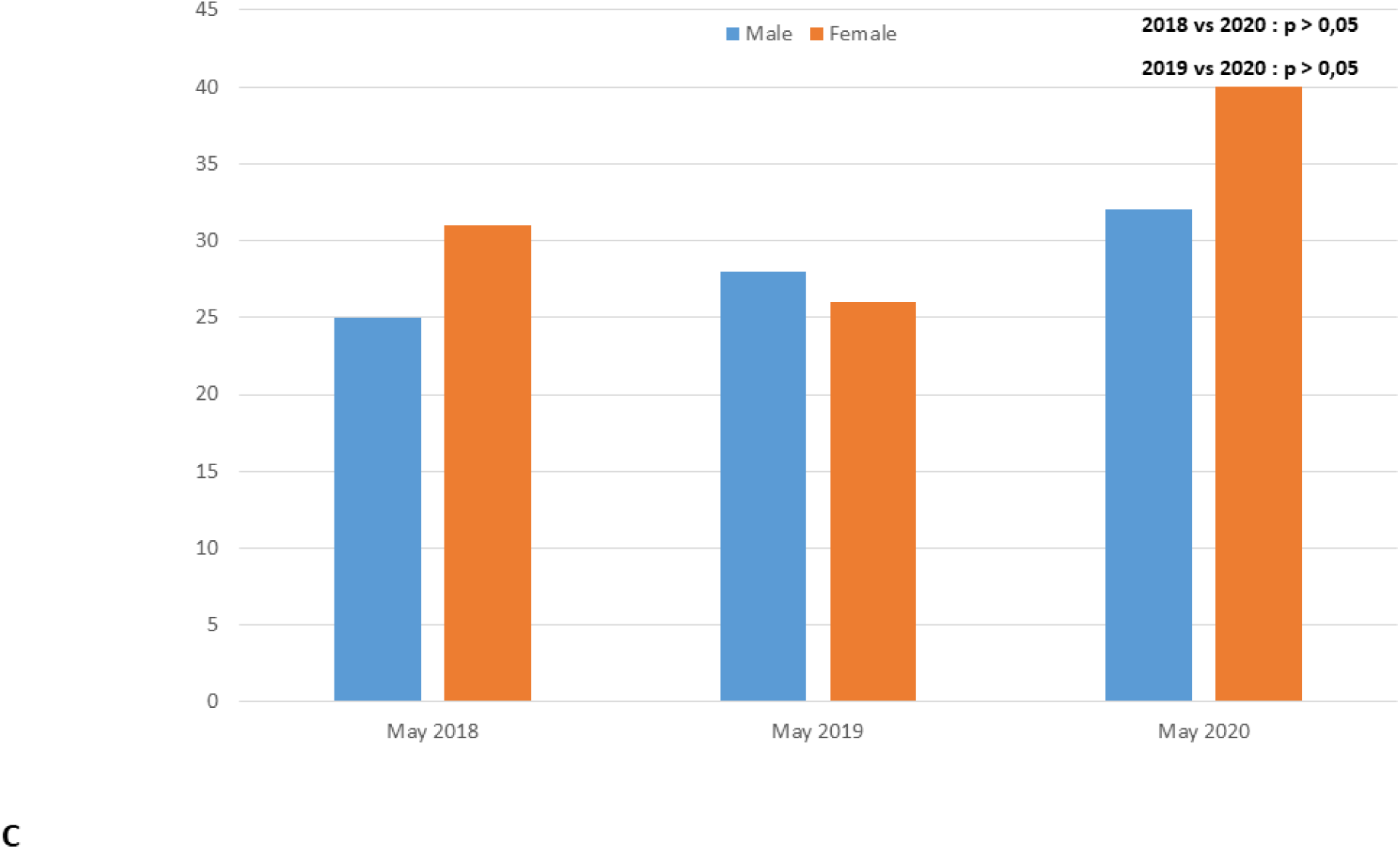

**Figure 4.**
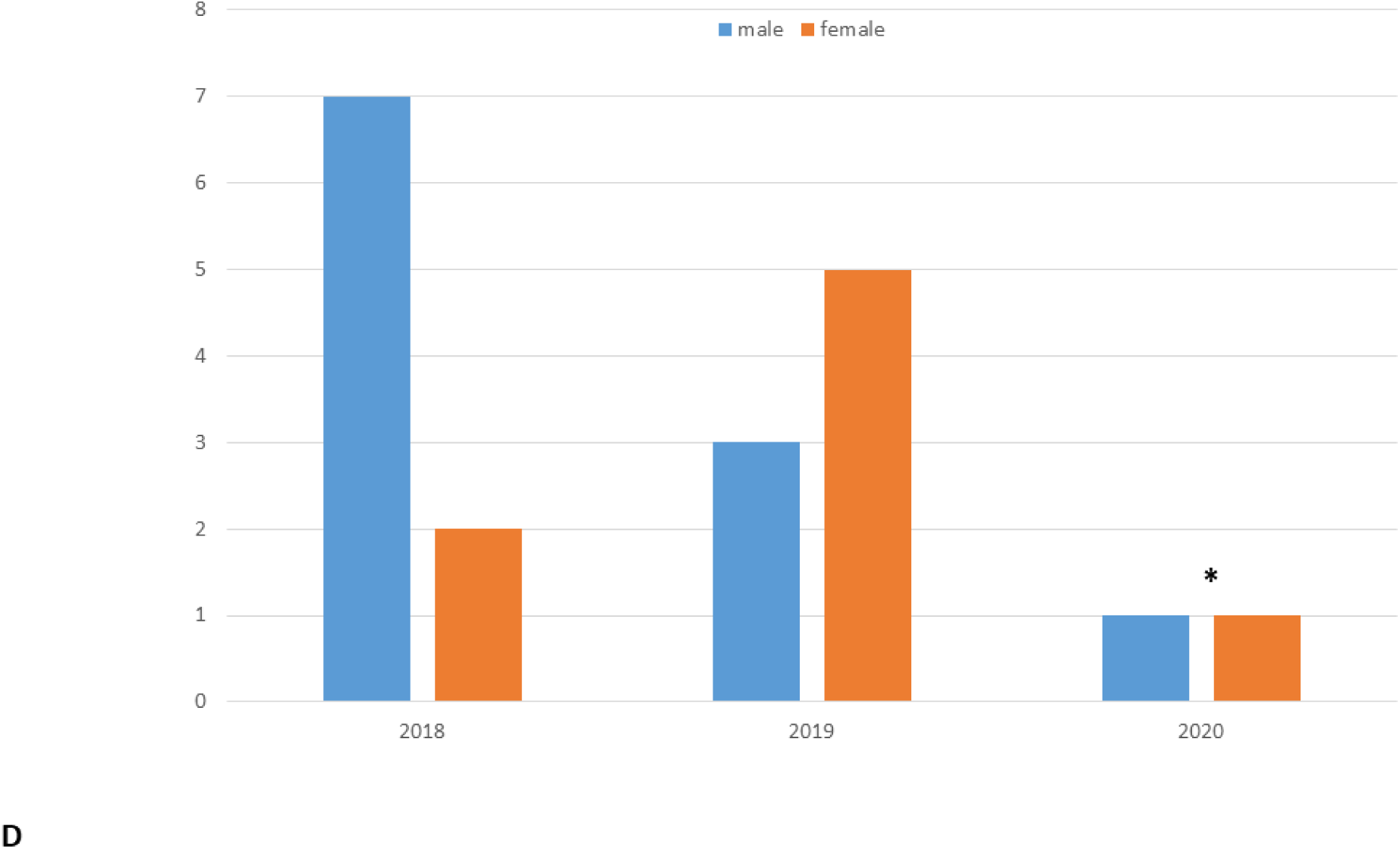

## Discussion

Looking at the trend of Malaria diagnoses in children under five years of age along the three investigated years, the number of diagnoses did not change significantly during COVID-19. Although an initial drop of Malaria cases has been documented in April 2020, when Sierra Leone started the lockdown due to the first cases of COVID-19, we cannot exclude that it is a casual fluctuation in Malaria diagnoses. Interestingly, when the lockdown was retired at the beginning of May 2020, a sudden increase in Malaria diagnoses have been registered. Again, a causal relationship with the reduction of restrictive measures related to COVID-19 cannot be demonstrate, but not even excluded.

According to local doctors and community health workers, in Sierra Leone health authorities and healthcare workers worked hard to ensure that people would not miss malaria care as happened during EVD outbreak. In fact, despite at the beginning of April 2020 local health workers reported an initial reduction on malaria diagnosis, mainly due to fear of contracting COVID-19 in the health centers, after a number of health education activities people came back to the health facilities when needed. In fact, on May 2020, when COVID-19 cases were still increasing in the country, the number of malaria diagnoses in our center in children under 5 years of age raised as well. To achieve this result, the Sierra Leone government took preventive strategies after the lessons learned during EVD outbreak. In particular:

- Education and information strategies aimed to eliminate the perception that health staff is infecting people with COVID-19
- Continuous health education on the use of insecticide treated net which has helped greatly in the reduction of malaria positive cases
- Implementation of Malaria intermittent preventive treatment in infants (IPTi) given during the routine immunization activities to infants aged 10 weeks to 9 months
- nationwide free distribution of ITNs given to every household in May 2020, while this campaign has been interrupted during the previous Ebola outbreak
- systematic malaria diagnostics as part of fever management and measures for early detection and treatment of malaria, including presumptive malaria treatment
- Implementation of community-based health workers for social engagement and monitoring of peripheries
- Continuation of malaria drug and test supply

This experience in a local health center previously involved by the EVD shows how a pro-active approach is necessary in order to keep appropriate care for major killers in Africa, performing proactive screening not only to diagnose COVID-19 but also Malaria. This is a necessary step in sub-Saharan Africa since a missed diagnosis of malaria bear significant public health consequences for the community allowing the further spread of Malaria, similarly to a missed diagnosis of COVID-19 (12). The clinical presentation of malaria is similar to COVID-19; moreover, both can be asymptomatic or pauci-symptomatic or even present with systemic critical disease (12). This further emphasizes the need to actively look for both diagnoses in malaria endemic countries.

We are aware that our report has several limitations to address. The retrospective nature is a limitation itself. Moreover, we have been able to collect absolute numbers, and no comprehensive epidemiological/demographic data from the health center are currently available, since the pandemic and the related restrictions are creating a higher workload for local workers and, at the same time, limits their possibility of interacting with other offices. Moreover, the restrictive measures lead to local organizational changes, including a reduction of the number of health workers concomitantly working at each health post, in order to reduce the risk of contagion of local workers, already extremely low in the area. Therefore, there were no enough human resources, and time, to allow us a timely and more detailed data collection. Also, ecological changes during the three study periods may have potentially influenced the trend of local malaria diagnoses, although currently there are no published evidences of unusually different rainy or dry seasons in the study period. Last, our data reflects the results of a national campaign in a single health center in Sierra Leone, therefore these data cannot be generalized to the whole country or, in general, to Sub-Saharan Africa.

Although some difficulties in keeping effectively active Tuberculosis centers in sub-Saharan Africa has been already documented in Sierra Leone (4), in the case of Malaria in young children, the national health system has been apparently more successful, at least in the initial phases of the pandemic, since after an initial reduction of new diagnoses in April (first month of local lockdown), on May 2020 a raise of new cases has been registered. Obviously, the management of chronic conditions could be considered more difficult, as direct-observed therapies require multiple evaluations that during lockdown are not easily allowed, and telemedicine services are not yet implemented in low-income countries. However, our study, although limited by its retrospective nature, can suggests that a proactive approach in the management of malaria in endemic countries during COVID-19, even including educational activities to sensitize the local population, is important and may be potentially useful in order to guarantee successful malaria diagnosis and treatment. However, long-term observational studies including more centers, and particularly the peripheral health centers of low-income countries, are needed to understand if a proactive and educational approach will be sufficient to prevent excess of malaria deaths due to disruption of the local health systems related to the SARS-CoV-2 pandemic.

## Data Availability

available upon request

## Acknowledgments

We are grateful to all our colleagues that supported the development of local health services and training of community health workers in the Western Rural Area of Sierra Leone and, in particular, Ismail Jaber, Matilda Yamba, Prince Williams, Memunatu N Kallon, Nee Turay, Pietro Sollena, Alessia De Nisco, Stefano Rocchi, Irene Sollena, Francesca Vassallo, Vittorio Sabatino, Mara Caramia, Gregory Leeb, Valeria Pansini, Filippo Bruno, Andrea Deidda, Arianna Cafarotti, Davide Guglielmi, Maria Giulia Conti, Jessica Balerna, Francesco Madeddu, Mattia Belardinelli, Daniele Barbuto, Luigi Torricelli, Rachel Mannings, Surf4Children Onlus

We are grateful to the Research Center of Global Health of the Istituto di Sanità Pubblica, Università Cattolica del Sacro Cuore, Roma, Italia, for the scientific and cultural support in this research project.

We are also grateful to Cassa Galeno, which granted Danilo Buonsenso a project aimed at the development of an Italian-Sierra Leone research network in pediatrics.

## Figure legend

*P. falciparum* malaria diagnoses in children under five years of age evaluated in a health center in Sierra Leone, during the period January-May 2018, 2019 and 2020 (a). Details of Malaria diagnosis in April 2018, 2019 and 2020 (b) and in May 2018, 2019 and 2020 (c). Malaria deaths during the study period (d)

## Notes

**Conflict of interest statement** The authors declare that the research was conducted in the absence of any commercial or financial relationships that could be construed as a potential conflict of interest

### Competing Interest Statement

The authors have declared no competing interest.

### Author Declarations

The study was approved by a local commission composed of the responsible of the Malaria unit of the local health centre (S.S.), the research team of the Bureh Town Community Hospital of Sierra Leone, the headman of the community and the old men of the village, in a similar way to what happens for all the important political and economic decisions in the examined area (n25_may-2020).

